# Health Systems Disparities and Publicly Funded Health Insurance Outcomes in India: Insights from Comprehensive Annual Modular Survey, 2022-23

**DOI:** 10.1101/2025.03.14.25323951

**Authors:** Sekhar Bonu, Indu Bhushan

## Abstract

India’s health systems vary significantly across its states and Union Territories due to institutional, social, economic and historical factors. Publicly funded health insurance schemes (PFHI), mainly Ayushman Bharat-Prime Minister Jan Aarogya Yojana (AB-PMJAY) introduced in 2018 and state health insurance schemes introduced over the past two decades, aim to improve access to healthcare and reduce financial burden. This study examines the relationship between the diverse state health systems across 36 states and union territories (UTs) of India and PFHI outcomes using the Comprehensive Annual Modular Survey, 2022–23 data covering over 302,000 households. States and UTs are classified as high or low health systems performers based on the state health index (representing the capability of primary healthcare), the district hospital index (indicating the strength of public district hospitals), and the state’s prior experience with health insurance before the launch of AB-PMJAY (representing implementation experience).

The study results reveal that states with better health systems show higher PFHI coverage. A higher state health index is associated with lower outpatient out-of-pocket expenditure (OOPE) (coefficient - 0.100, p<0.01). A higher district hospital index is associated with lower inpatient OOPE (coefficient - 0.297, p<0.01). Prior insurance experience is associated with higher inpatient utilisation. In addition to its independent effect on OOPE, better district hospitals appear to improve the efficacy of PFHI in achieving its intended outcomes. The interaction terms between PFHI and the district hospital index indicate higher inpatient utilisation (adjusted odds ratio (AOR) 1.164, p<0.01), lower inpatient catastrophic health expenditure (CHE) (AOR 0.883, p<0.05) and lower inpatient OOPE (coefficient - 0.252, p < 0.01).

The findings support strengthening primary health care and public hospital systems, especially in low-performing states, to amplify the benefits of PFHI. The study also supports expanding PFHI to cover outpatient care. By aligning PFHI with reliable public health system improvements, India can enhance healthcare access and equity, increase financial protection, and achieve universal health coverage.

**What is already known on this topic:** As reflected in the state health index and district hospital index, India’s health system is diverse and influenced by social, economic, historical and institutional factors. Publicly funded health insurance (PFHI) schemes aim to improve healthcare access and reduce financial burdens.

**What this study adds:** Findings reveal that states with better health systems are associated with higher PFHI coverage, higher inpatient care utilisation, and better inpatient care financial protection, among others.

**How this study might affect research, practice, or policy:** Policymakers can use insights from the study to prioritise investments in primary health care and district hospital infrastructure, particularly in low-performing states, to amplify PFHI outcomes.

## 1. Introduction

The health systems in India, including primary health care and district public hospitals, vary significantly across the 36 states and Union Territories (UTs).(1,2) Various state governments have introduced publicly funded health insurance (PFHI) schemes over the past two decades (Table 1), complementing the Union government’s efforts through the Rashtriya Swasthya Bima Yojana, introduced in 2008, which was replaced with Ayushman Bharat-Prime Minister Jan Aarogya Yojana (AB-PMJAY) in 2018. (3,4) The interplay between India’s diverse state government-led health systems and the phased introduction of health insurance lends itself to a natural experiment to study the association between the strength of health systems and health insurance outcomes. (4)

**Tables 1:**
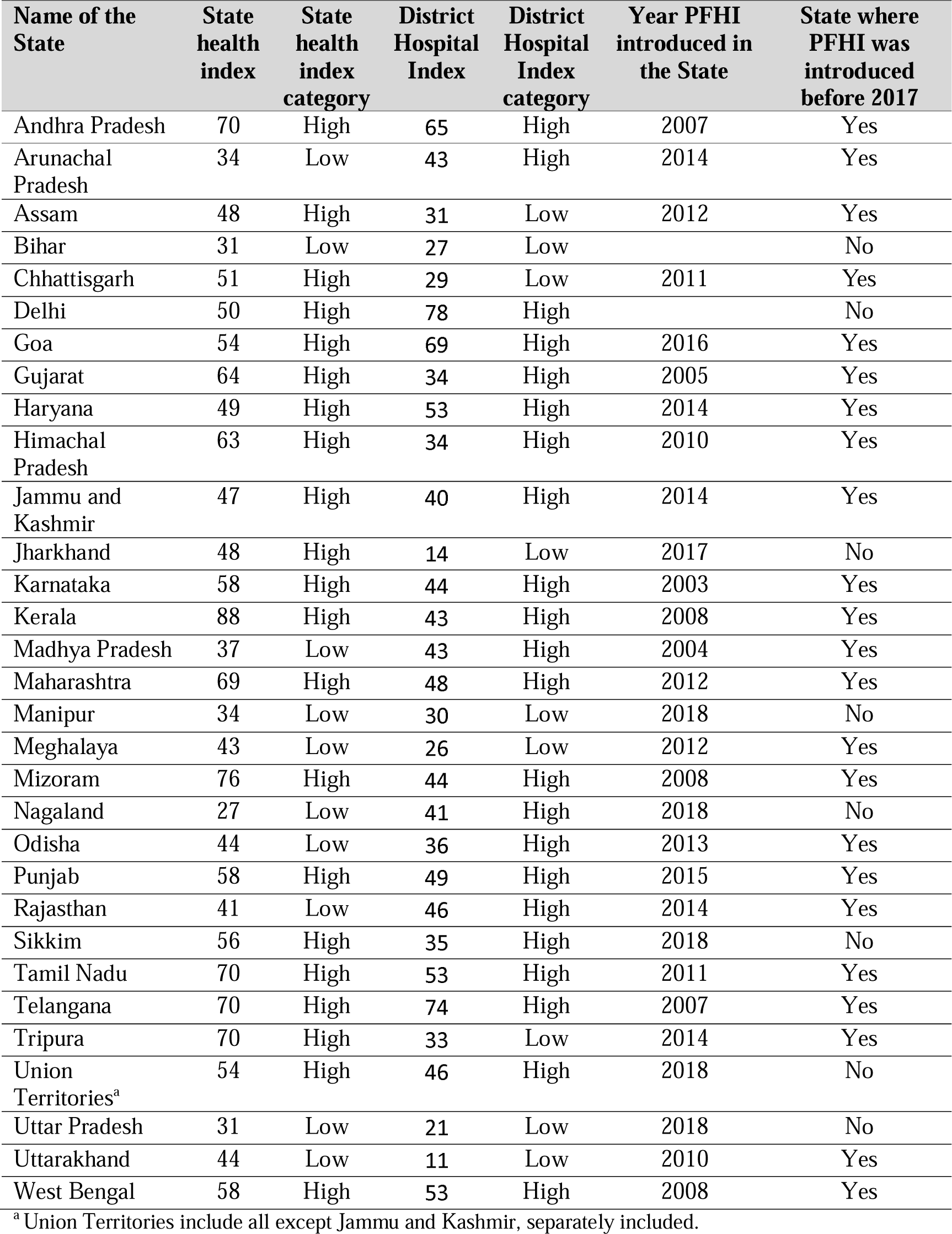
State health index, Hospital Index, Year PFHI first launched, and details of health insurance history.

| Name of the State | State health index | State health index category | District Hospital Index | District Hospital Index category | Year PFHI introduced in the State | State where PFHI was introduced before 2017 |
| --- | --- | --- | --- | --- | --- | --- |
| Andhra Pradesh | 70 | High | 65 | High | 2007 | Yes |
| Arunachal Pradesh | 34 | Low | 43 | High | 2014 | Yes |
| Assam | 48 | High | 31 | Low | 2012 | Yes |
| Bihar | 31 | Low | 27 | Low |  | No |
| Chhattisgarh | 51 | High | 29 | Low | 2011 | Yes |
| Delhi | 50 | High | 78 | High |  | No |
| Goa | 54 | High | 69 | High | 2016 | Yes |
| Gujarat | 64 | High | 34 | High | 2005 | Yes |
| Haryana | 49 | High | 53 | High | 2014 | Yes |
| Himachal Pradesh | 63 | High | 34 | High | 2010 | Yes |
| Jammu and Kashmir | 47 | High | 40 | High | 2014 | Yes |
| Jharkhand | 48 | High | 14 | Low | 2017 | No |
| Karnataka | 58 | High | 44 | High | 2003 | Yes |
| Kerala | 88 | High | 43 | High | 2008 | Yes |
| Madhya Pradesh | 37 | Low | 43 | High | 2004 | Yes |
| Maharashtra | 69 | High | 48 | High | 2012 | Yes |
| Manipur | 34 | Low | 30 | Low | 2018 | No |
| Meghalaya | 43 | Low | 26 | Low | 2012 | Yes |
| Mizoram | 76 | High | 44 | High | 2008 | Yes |
| Nagaland | 27 | Low | 41 | High | 2018 | No |
| Odisha | 44 | Low | 36 | High | 2013 | Yes |
| Punjab | 58 | High | 49 | High | 2015 | Yes |
| Rajasthan | 41 | Low | 46 | High | 2014 | Yes |
| Sikkim | 56 | High | 35 | High | 2018 | No |
| Tamil Nadu | 70 | High | 53 | High | 2011 | Yes |
| Telangana | 70 | High | 74 | High | 2007 | Yes |
| Tripura | 70 | High | 33 | Low | 2014 | Yes |
| Union Territories<sup>a</sup> | 54 | High | 46 | High | 2018 | No |
| Uttar Pradesh | 31 | Low | 21 | Low | 2018 | No |
| Uttarakhand | 44 | Low | 11 | Low | 2010 | Yes |
| West Bengal | 58 | High | 53 | High | 2008 | Yes |

<sup>a</sup> Union Territories include all except Jammu and Kashmir, separately included.

The Indian healthcare system is diverse, complex, and changing, with stubborn challenges.(5) Due to historical, social, cultural, and economic reasons, states in India vary widely in social determinants of health and healthcare-related inputs, processes, and outputs. (6–10) The health systems in India are primarily state-government-led, operating within India’s federal framework. (11,12) The Union government and state governments finance health care; however, many state governments depend on the Union government’s fiscal transfers and schemes to bolster their healthcare budgets. (7,13) These have led to significant state-level and district-level variations in health systems and universal health coverage (UHC). (8,11,14) NITI Aayog, the Government of India’s apex think tank, introduced and acknowledged significant variation in the state health index, ranging from as low as 31 in Bihar to 88 in Kerala. (1) Similarly, NITI Aayog found significant hospital system variations in 707 district hospitals covering most of India. (2)

State-level variations in inpatient care and out-of-pocket expenditures (OOPE) are well established. (15–18) India’s health system is mixed: 70% of outpatient and 58% of inpatient services are through for-profit or not-for-profit providers. (3) Hospital bed availability varies from 35 per 10,000 in Tamil Nadu to as low as 2.45 in Bihar, and life expectancy varies by 10 years between high-achiever Kerala and the bottom states. (3) Many states have procurement and supply issues for public hospitals, with some exceptions where strong institutions ensure efficient procurement and regular supplies of medicines and diagnostics.(3)

Health insurance coverage for inpatient care increased substantially after the launch of AB-PMJAY, aided by prior state-level initiatives in some states. (4,9) These reforms reflect an increasing focus on financial risk protection for achieving UHC in India. (16) The National Rural Health Mission (NRHM), launched in 2005, has made progress in strengthening primary health care, albeit unevenly across different states. (19) India’s healthcare challenges include a lack of a comprehensive systems approach to health policies, inadequate and inefficient health financing mechanisms, and fragmentation between the public and private sectors. (20)

In this broad and diverse context, the study explores whether states with better primary health care, district public hospital systems, and prior experience with health insurance programs are associated with better access and outcomes of PFHI. To achieve its objectives, the study uses the natural experiment unravelling in India resulting from the phased introduction of health insurance and divergent health systems. The study findings can provide policymakers in India and the developing world insights for efficient resource allocation between the growing demand for the PFHI budget and strengthening core primary health care and public hospital systems, mainly where health care depends on a mix of public and private provision.

PFHI covers only inpatient care in India. A well-established primary health care network is expected to improve outpatient care and reduce related OOPE and CHE. PFHI and better public district hospitals are expected to increase inpatient care and reduce related OOPE and CHE. District hospitals also establish benchmarks for the private sector as the quality of private hospitals is seen to be closely related to that of public hospitals. (21,22)

At the time of AB-PMJAY’s launch, several states already had existing health insurance schemes or were actively implementing RSBY. This prior experience gave them a distinct advantage, as they had established implementation agencies, a greater awareness about the scheme among the target population, and an existing network of hospitals offering services. In contrast, other states faced additional challenges in setting up the scheme and building momentum for its implementation.

This study examines the following questions: Are strong state primary health care systems associated with higher PFHI enrollments, more outpatient use, and lower OOPE and CHE during outpatient care? Are reliable district hospitals associated with higher PFHI enrollments, higher inpatient use, and lower OOPE and CHE during inpatient care? Does longer experience implementing PFHI lead to better use of inpatient services and lower inpatient OOPE and CHE?

## 2. Methods

### Data

This study uses data from the Comprehensive Annual Modular Survey (CAMS) 2022-23 (79^th^ National Sample Survey round), conducted by the National Sample Survey Office from July 2022 to June 2023. (23) CAMS (2022–23) used a stratified multi-stage design to sample 173,096 rural and 128,990 urban households. The survey obtained information on whether any household members were covered under one or more health insurance schemes at the time of the survey, including PFHI schemes that include AB-PMJAY and State health insurance schemes and others (Employees’ State Insurance Scheme (ESIS), Central Government Health Schemes (CGHS), privately purchased commercial insurance, or any other employer-provided health insurance schemes).

Data were collected on whether any household member sought treatment through outpatient care (that includes all non-hospitalised care, including preventive measures and self-care) within the past 30 days of the survey along with associated OOPE and inpatient care (including hospitalisation and institutional deliveries) within the past 365 days of the survey, along with corresponding OOPE. Medical expenditures covered packages and non-package components, such as doctor’s fees, medicines, diagnostic tests, and other medical costs. Details were also obtained regarding the amount reimbursed by medical insurance providers or employers for outpatient and inpatient care.

### Outcome Variables

PFHI aims to ensure that beneficiaries access quality healthcare services without financial distress. PFHI schemes in India cascade through several steps, including enrolment, awareness about enrolment and associated benefits, access to and selection of participating healthcare providers, and use of services. (24) Within this broad framework, this study seeks to assess the association of proxies of the strength of health systems on key health insurance-related outcomes in three broad categories of outcome variables: health insurance coverage, healthcare use and financial protection during healthcare episodes. The first category focuses on the enrollment of households in any insurance. The second outcome is about the use of outpatient and inpatient care. The third category examines the level of financial protection by assessing OOPE and CHE. The outcomes used in the study are as follows:

a. Any health insurance coverage (binary: 1 = yes, 0 = no): whether any household member has at least one health insurance coverage.
b. Health care use (binary: 1 = yes, 0 = no): (a) use of outpatient care in the last 30 days before the survey; and (b) use of inpatient care in the last 365 days before the survey.
c. CHE (binary: 1 = yes, 0 = no): whether households incur CHE while seeking health care where CHE is defined as health expenditure of more than 10% of household monthly consumption expenditure for outpatient care and 10% of annual consumption expenditure for inpatient care.
d. OOPE (continuous variable, Indian rupees (INR) spent): Expenditure incurred by households while seeking outpatient or inpatient care after adjusting to any reimbursements by health insurance agencies.

### Independent Variables

The independent variables include proxies to health systems, which are the exposure variables that are of interest to the study, and socio-economic covariates as control variables.

### Health systems

Three independent variables are deployed as proxies for (a) the state health index and (b) the district hospital index, (c) health insurance coverage, and (d) prior PFHI experience for a state.

a. **State health index:** The study uses a categorical variable (low and high-performing states) constructed using the fourth round of the state health index prepared jointly by the NITI Aayog, the apex think tank of the government of India, the Ministry of Health and Family Welfare, and the World Bank. (1) The index comprises 12 health outcomes, four related to governance and information domains and 27 indicators related to inputs and processes related to primary health care. (25) The index highlights the effectiveness of state primary healthcare infrastructure and governance.(26) Although a study has raised questions about the index, it is by far the best publicly available data to use as a proxy for the strength of primary care at the state level. States with scores equal to or less than 44 are categorised as “0” and those above 44 as “1” (Table 1).
b. **District hospital index**: The study uses categorical variables (low and high-performing states) built using the first comprehensive assessment of the district hospitals in India. (2) The NITI Aayog assessment of India’s district hospitals utilised ten key performance indicators, including infrastructure (e.g., beds, doctors, nurses, paramedics, diagnostic and healthcare facilities) and output indicators (e.g., caesarean section surgery rates and bed occupancy). The study involved 707 district hospitals from 36 States and UTs, with data from the 2017–18 financial year validated through physical records by the National Accreditation Board for Hospitals and Healthcare Providers. States with a district hospital index equal to or below 33 are scored “0”, and those above are scored “1” (Table 1).^1^
c. **Health Insurance Coverage:** Health insurance coverage uses two dummy variables: 0=uninsured, 1=PFHI schemes, including AB-PMJAY and those funded by state governments, and 0=uninsured and 1=other insurance (ESIC/CGHS/and other employer-provided insurance and private insurance).
d. **Prior Health Insurance Coverage:** States with state-level PFHI experience prior to the launch of AB-PMAY in 2017 are categorised as 1, and states that rolled out health insurance as a result of AB-PMJAY or other state-level initiatives after 2017 are categorised as ‘0’. While many states are categorised as “1”, the following states and all UTs are categorised as “0”: Bihar, Delhi, Jharkhand, Manipur, Nagaland, Sikkim, UTs, and Uttar Pradesh (Table 1).

### Other Covariates

The other independent variables are designed to capture socio-economic, demographic, and regional disparities. Rural/urban residence indicates whether a household is rural or urban. Social groups include Scheduled Tribes, Scheduled Castes, Other Backward Classes, and Others. Religion is a categorical variable, including Hindu, Muslim, Christian, and Others. Households are divided into expenditure deciles based on the household’s monthly per capita expenditure, a proxy for income. The education of the household head is categorised as illiterate or less than primary, primary, middle, secondary/senior secondary, and graduate and above.

### Statistical analysis

Descriptive statistics are used to profile sampled households, PFHI coverage, and use of outpatient and inpatient care (Tables 2 and 3). Cross-tabulations examined the association between independent variables and CHE and OOPE (Table 3). Five logistic regression models attempt to quantify the relationship between health systems and health insurance coverage, as well as the use of healthcare and CHE, after controlling for various covariates in the model. To investigate the relationship between health systems and OOPE, two Heckman’s two-stage selection models were used to address potential sample selection bias. OOPE data are log-transformed to handle skewed distributions commonly observed in such data.

**Table 2:**
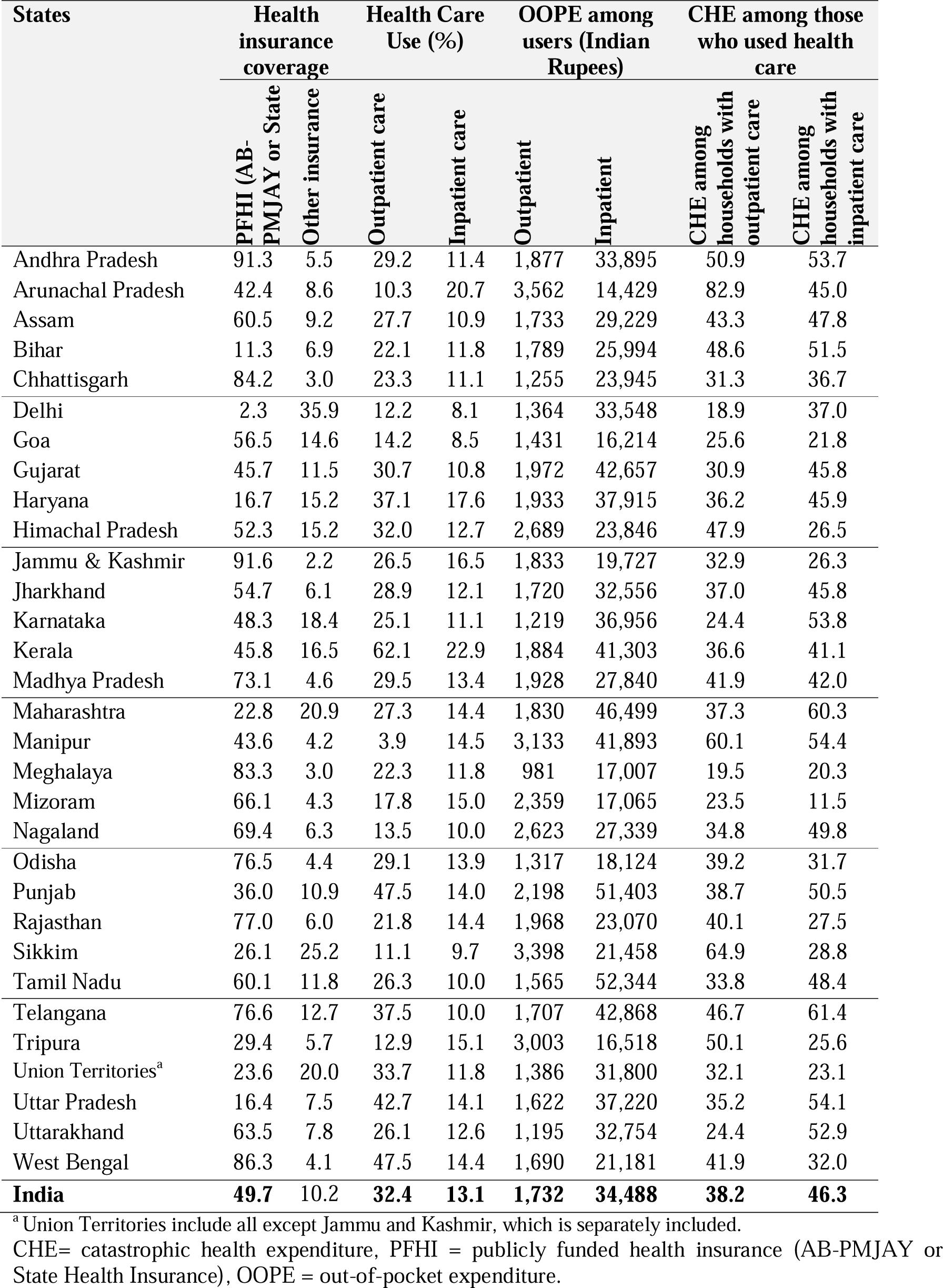
Variation in health insurance coverage, health care use, out-of-pocket expenditure (OOPE), and catastrophic health expenditure (CHE) among States and Union Territories of India.

| States | Health insurance coverage |  | Health Care Use (%) |  | OOPE among users (Indian Rupees) |  | CHE among those who used health care |  |
| --- | --- | --- | --- | --- | --- | --- | --- | --- |
|  | PFHI (AB-PMJAY or State | Other insurance | Outpatient care | Inpatient care | Outpatient | Inpatient | CHE among households with outpatient care | CHE among households with inpatient care |
| Andhra Pradesh | 91.3 | 5.5 | 29.2 | 11.4 | 1,877 | 33,895 | 50.9 | 53.7 |
| Arunachal Pradesh | 42.4 | 8.6 | 10.3 | 20.7 | 3,562 | 14,429 | 82.9 | 45.0 |
| Assam | 60.5 | 9.2 | 27.7 | 10.9 | 1,733 | 29,229 | 43.3 | 47.8 |
| Bihar | 11.3 | 6.9 | 22.1 | 11.8 | 1,789 | 25,994 | 48.6 | 51.5 |
| Chhattisgarh | 84.2 | 3.0 | 23.3 | 11.1 | 1,255 | 23,945 | 31.3 | 36.7 |
| Delhi | 2.3 | 35.9 | 12.2 | 8.1 | 1,364 | 33,548 | 18.9 | 37.0 |
| Goa | 56.5 | 14.6 | 14.2 | 8.5 | 1,431 | 16,214 | 25.6 | 21.8 |
| Gujarat | 45.7 | 11.5 | 30.7 | 10.8 | 1,972 | 42,657 | 30.9 | 45.8 |
| Haryana | 16.7 | 15.2 | 37.1 | 17.6 | 1,933 | 37,915 | 36.2 | 45.9 |
| Himachal Pradesh | 52.3 | 15.2 | 32.0 | 12.7 | 2,689 | 23,846 | 47.9 | 26.5 |
| Jammu & Kashmir | 91.6 | 2.2 | 26.5 | 16.5 | 1,833 | 19,727 | 32.9 | 26.3 |
| Jharkhand | 54.7 | 6.1 | 28.9 | 12.1 | 1,720 | 32,556 | 37.0 | 45.8 |
| Karnataka | 48.3 | 18.4 | 25.1 | 11.1 | 1,219 | 36,956 | 24.4 | 53.8 |
| Kerala | 45.8 | 16.5 | 62.1 | 22.9 | 1,884 | 41,303 | 36.6 | 41.1 |
| Madhya Pradesh | 73.1 | 4.6 | 29.5 | 13.4 | 1,928 | 27,840 | 41.9 | 42.0 |
| Maharashtra | 22.8 | 20.9 | 27.3 | 14.4 | 1,830 | 46,499 | 37.3 | 60.3 |
| Manipur | 43.6 | 4.2 | 3.9 | 14.5 | 3,133 | 41,893 | 60.1 | 54.4 |
| Meghalaya | 83.3 | 3.0 | 22.3 | 11.8 | 981 | 17,007 | 19.5 | 20.3 |
| Mizoram | 66.1 | 4.3 | 17.8 | 15.0 | 2,359 | 17,065 | 23.5 | 11.5 |
| Nagaland | 69.4 | 6.3 | 13.5 | 10.0 | 2,623 | 27,339 | 34.8 | 49.8 |
| Odisha | 76.5 | 4.4 | 29.1 | 13.9 | 1,317 | 18,124 | 39.2 | 31.7 |
| Punjab | 36.0 | 10.9 | 47.5 | 14.0 | 2,198 | 51,403 | 38.7 | 50.5 |
| Rajasthan | 77.0 | 6.0 | 21.8 | 14.4 | 1,968 | 23,070 | 40.1 | 27.5 |
| Sikkim | 26.1 | 25.2 | 11.1 | 9.7 | 3,398 | 21,458 | 64.9 | 28.8 |
| Tamil Nadu | 60.1 | 11.8 | 26.3 | 10.0 | 1,565 | 52,344 | 33.8 | 48.4 |
| Telangana | 76.6 | 12.7 | 37.5 | 10.0 | 1,707 | 42,868 | 46.7 | 61.4 |
| Tripura | 29.4 | 5.7 | 12.9 | 15.1 | 3,003 | 16,518 | 50.1 | 25.6 |
| Union Territories<sup>a</sup> | 23.6 | 20.0 | 33.7 | 11.8 | 1,386 | 31,800 | 32.1 | 23.1 |
| Uttar Pradesh | 16.4 | 7.5 | 42.7 | 14.1 | 1,622 | 37,220 | 35.2 | 54.1 |
| Uttarakhand | 63.5 | 7.8 | 26.1 | 12.6 | 1,195 | 32,754 | 24.4 | 52.9 |
| West Bengal | 86.3 | 4.1 | 47.5 | 14.4 | 1,690 | 21,181 | 41.9 | 32.0 |
| **India** | **49.7** | **10.2** | **32.4** | **13.1** | **1,732** | **34,488** | **38.2** | **46.3** |

<sup>a</sup> Union Territories include all except Jammu and Kashmir, which is separately included.
CHE= catastrophic health expenditure, PFHI = publicly funded health insurance (AB-PMJAY or State Health Insurance), OOPE = out-of-pocket expenditure.

**Table 3:**
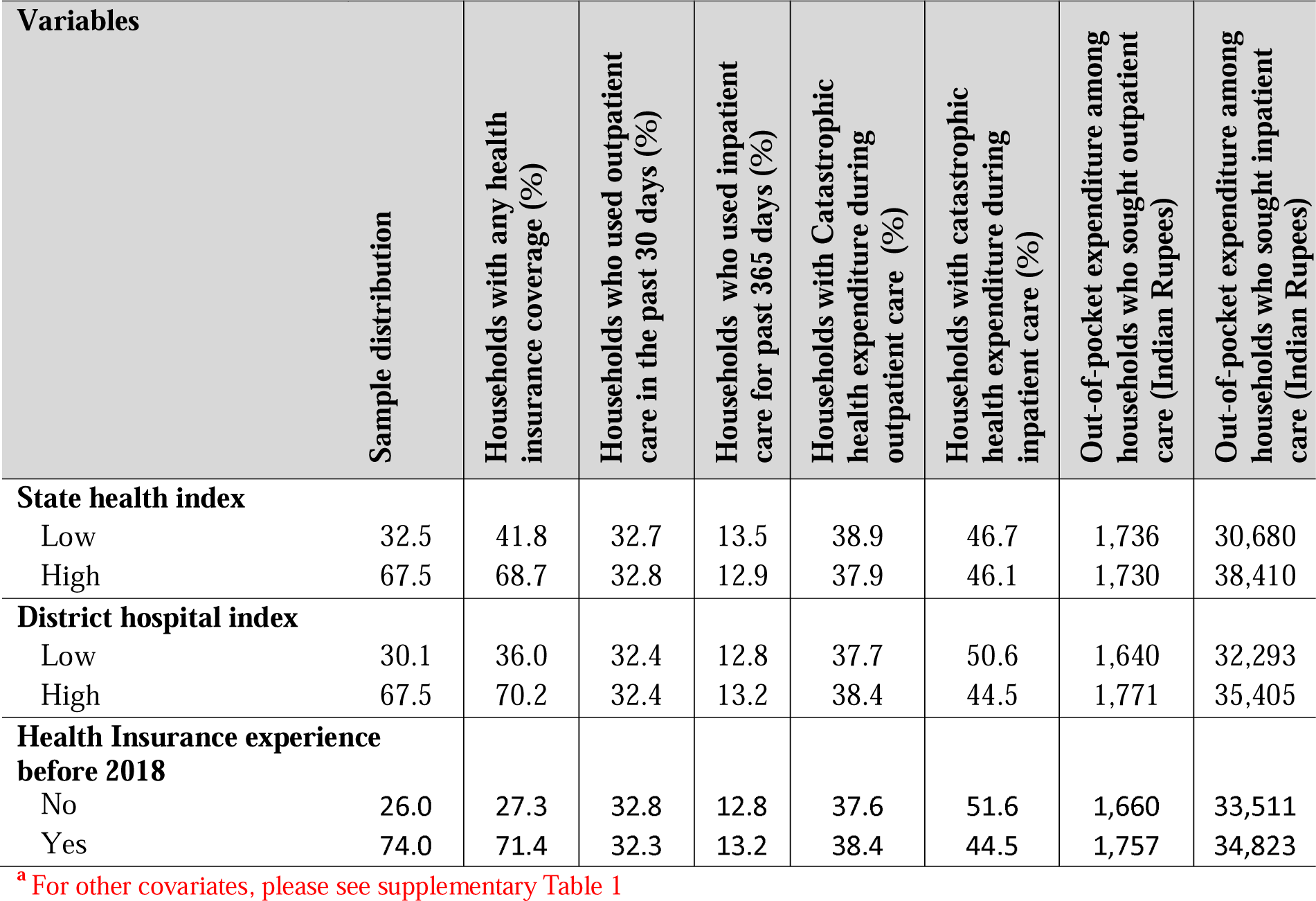
Sample distribution and bivariate estimates of outcome by study covariates^a^.

PFHI covers mainly inpatient care in India. Therefore, models relating to outpatients only model the state health index without the term’s interaction with PFHI. Those modelling inpatient care (Models 5 and 7) include the district hospital index and the interaction term between PFHI and the district hospital index.

Logistic Regression is used to model binary outcome variables: (a) health insurance coverage (Model 1) (Table 4), (b) outpatient care in the past 30 days (Model 2) (Table 4), (c) inpatient care in the past 365 days (Model 3) (Table 4), (d) CHE during outpatient care (Model 4), and (e) CHE during inpatient care (Model 5) (Table 5 for Models 4 and 5). In Models 1-5, outcome (Y_1_) is modelled as a binary outcome (1 if insured or zero otherwise, 1 if health care is used, or zero otherwise, or 1 if the households undergo CHE or zero otherwise. Robust standard errors are used to account for heteroscedasticity and ensure reliable estimates. The logistic regression equation is expressed as:

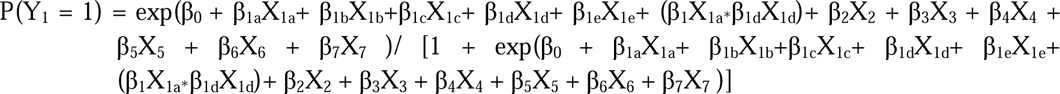

**Table 4:**
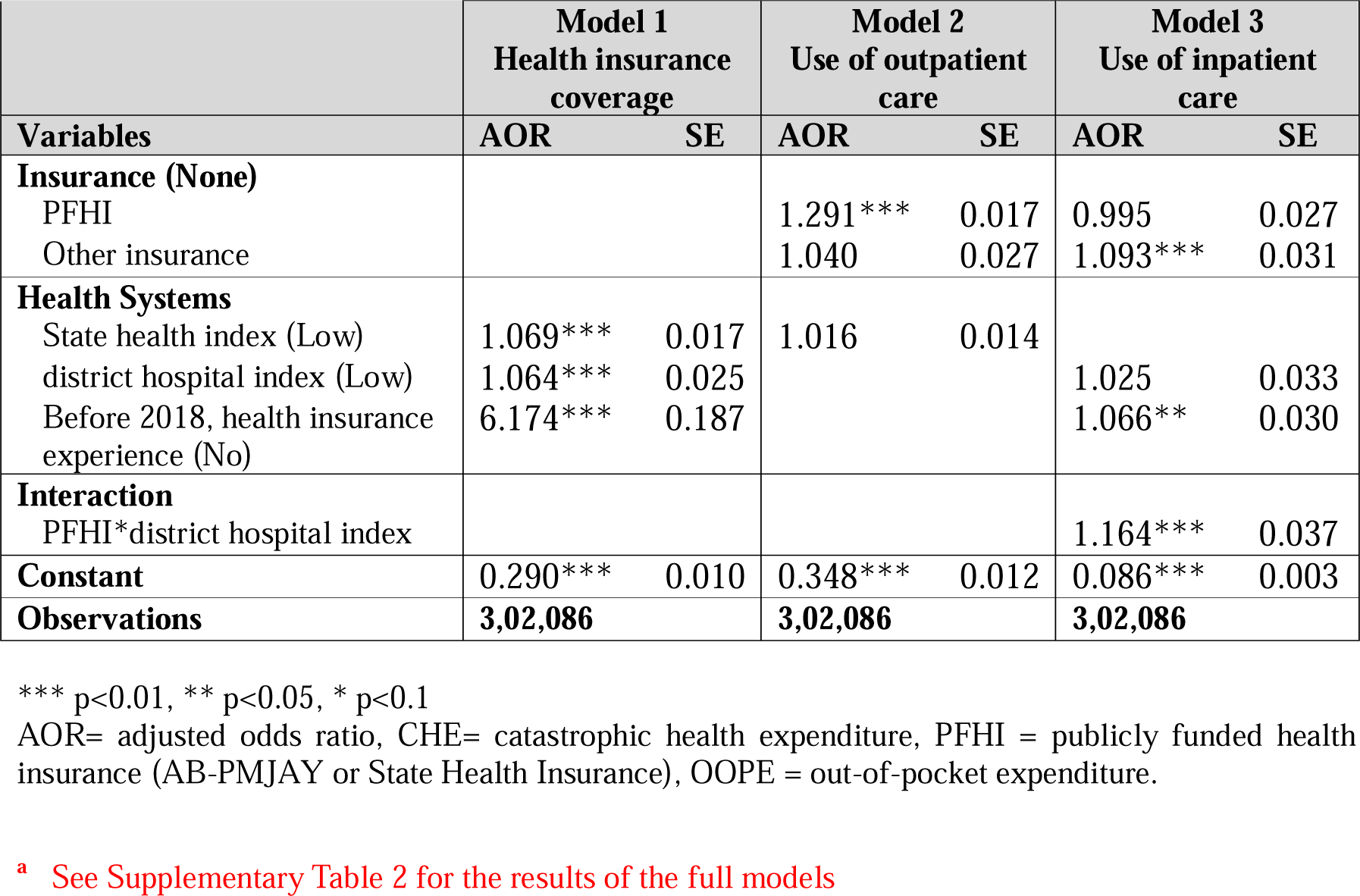
Logistic regression of (a) Health insurance coverage (Model 1), (b) Use of outpatient care (Model 2), and (c) Use of inpatient care (Model 3) ^a^.

**Table 5:**
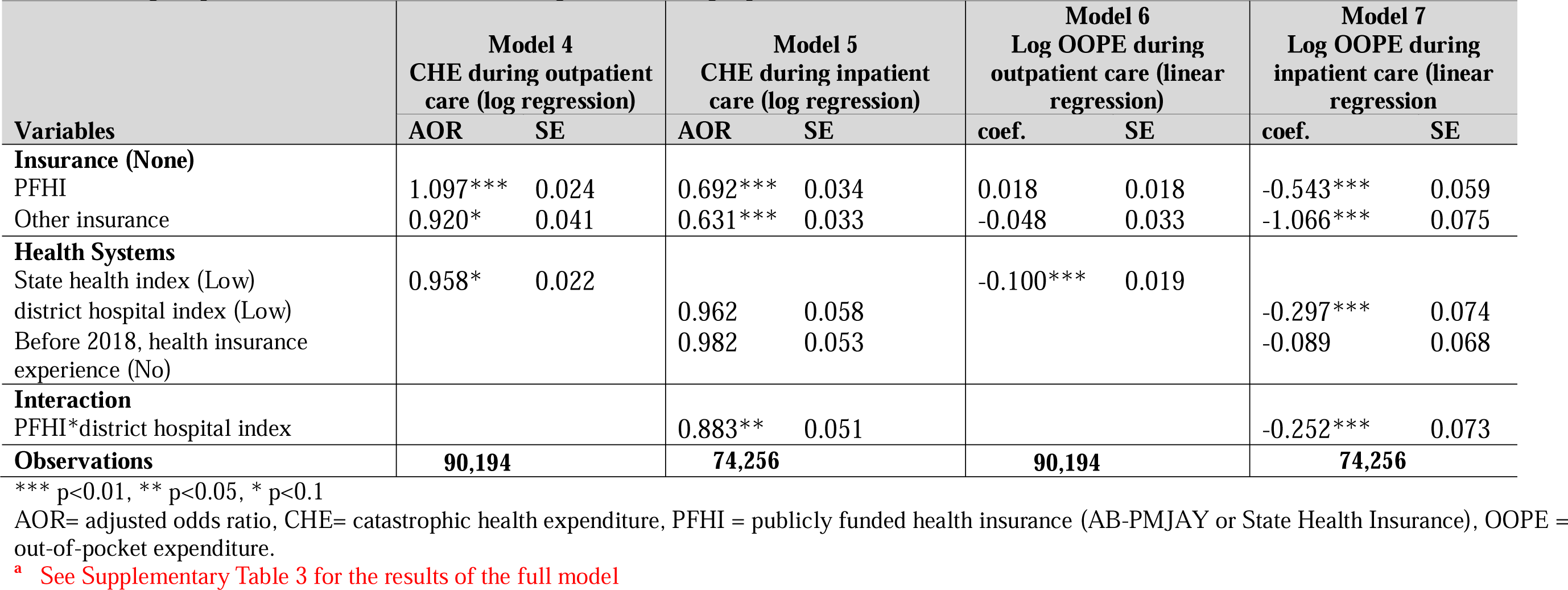
Logistic regression for (a) CHE during outpatient care (Model 4), (b) CHE during inpatient care (Model 5), and linear regression for (c) log OOPE during outpatient care (Model 6), and (d) Log OOPE during inpatient care ^a^.

Where:

- P(Y_1_ = 1) represents the probability of health care use or CHE
- β_0_ is the intercept, and β_1a_ to β_1e_ are the coefficient of special interest to the study relating to the health system. β_2_ to β_7_ are coefficients for the covariates that represent socioeconomic and demographic factors of the household.
- The health systems independent variables include: X_1a_: PFHI status (categorical); X_1b_: Other health insurance status (categorical); X_1c_: state health index (categorical, used only for outpatient care); X_1d_ district hospital index (categorical, used only for inpatient care); and X_1e_: Prior PFHI experience (categorical).
- Other covariates include: X_2_: Residence: urban/rural; X_3_: Religion (categorical); X_4_: Social group (categorical); X_5_: Household size (categorical); X_6_: Education of household head (categorical) and X_7_ Expenditure deciles (categorical).
- (β_1_X_1a*_β_1_X_1d_) is the coefficient of the interaction between PFHI and district hospital index used for Models 3,5 and 7 to assess the impact of PFHI where the district hospital index is better.

Statistical significance is assessed at conventional levels (p < 0.05, p < 0.01). Logistic regression output reports adjusted odds ratios (AOR) instead of coefficients explained by the equation.

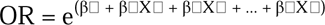

Heckman’s two-stage selection model uses a probit model, with the first stage estimating the probability of incurring any health expenditure and the second stage correcting for selection bias to provide unbiased and consistent estimates. Net OOPE was log-transformed to normalise the data, stabilise variance, and improve interpretability by allowing coefficients to be expressed in percentage terms. The selection equation (probit model) determines whether an individual incurs healthcare expenditure (binary outcome):

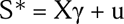

S = 1 if S* > 0 (households use health service either outpatient and/or hospitalised) or S = 0; otherwise. S represents the selection decision (whether the household uses health services), X includes the covariates, γ represents the coefficients, and *u* is the error term.

Outcome equation (expenditure model) conditional on incurring expenditure (S=1), the expenditure amount is modelled as:

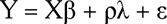

Where: - Y represents the log of healthcare expenditure (continuous outcome), X includes the covariates defined above, β represents the coefficients, λ is the Inverse Mills Ratio, derived from the selection equation to adjust for selection bias, ρ represents the coefficient of λ, capturing the correlation between errors in the selection and outcome equations, and ε is the error term.

## 3. Results Bivariate analysis

### State-level variations

Health insurance coverage in India exhibits considerable variation across states. Approximately 50% of the households self-reported that at least one member of the family was covered under PFHI, either PMJAY or state insurance programs, and notable gaps exist in states like Bihar (11.3%) and Uttar Pradesh (16.4%) (Table 2). In India, outpatient and inpatient care usage is 32.4% and 13.1%. States like Kerala (62.1%) and Punjab (47.5%) report significantly higher outpatient care utilisation, while states like Manipur (3.9%) and Tripura (12.9%) exhibit much lower rates. Inpatient care use is notably higher in states such as Kerala (22.9%) and Haryana (17.6%), contrasting with lower rates in Nagaland (10.0%) and Meghalaya (11.8%) (Table 2).

The average Out-of-Pocket Expenditure (OOPE) for outpatient care is Indian rupees (INR) 1,732, while inpatient care is significantly higher at INR 34,488. Gujarat reports the highest outpatient OOPE at INR 1,972, while Meghalaya has the lowest at INR 98. For inpatient care, Maharashtra (INR 46,499) and Kerala (INR 41,303) have the highest OOPE, whereas Odisha (INR 18,124) and Meghalaya (INR 17,007) report the lowest (Table 2).

The catastrophic health expenditure (CHE) for outpatient care is 38.2%. Meghalaya (19.5%) and Delhi (18.9%) report lower CHE percentages, while states like Andhra Pradesh (60.1%) and Arunachal Pradesh (82.9%) experience higher outpatient CHE (Table 2). The national average CHE for inpatient care stands at 46.3%. States such as Goa (21.8%) and Mizoram (11.5%) report lower CHE percentages, while Telangana (61.4%) and Maharashtra (60.3%) experience exceptionally high CHE (Table 2).

### Outcome variables with covariate of the study

Table 3 gives the “Sample Distribution” by various covariates.^2^ As for the state health index, 32.5% of households live in low-performing states, while 30.1% of households live in states with low district hospital index. States with health insurance experience before 2017 were 74% with prior experience. 67.1% of households reside in rural areas, 82.9% identify as Hindu, and heads lead 24.3% of households with less than primary education (See Supplementary Table 1 for other covariates).

The distribution of households with any health insurance coverage varies significantly by state health index, district hospital index, and prior insurance experience. Households in states with a high state health index report 68.7% insurance coverage, compared to 41.8% in states with a low index. Similarly, 70.2% of households have health insurance in states with a high district hospital index, whereas in states with a low hospital index, only 36% are covered. Additionally, households with health insurance experience before 2017 have a much higher coverage rate of 70.2%, compared to just 36% among those without prior experience (Table 3).

The distribution of households using outpatient and inpatient care shows notable differences based on state health index, district hospital index, and prior insurance experience. In states with a high state health index, 12.9% used inpatient care compared to 13.5% in households with a low state health index. For states with a high District hospital index, 13.2% used in-patient care compared to 12.8% for households with a low index. Households with prior insurance experience before 2017 showed the same outpatient care usage (32.4%) as those without experience. Inpatient care usage was high, 13.2% and 12.8%, respectively, with and without health experience prior to 2017 (Table 3).

CHE for outpatient and inpatient care exhibits variations across state health index, district hospital index, and prior insurance experience. For outpatient care, CHE is slightly lower in states with a high health index (37.9%) compared to low-index states (38.7%), while for inpatient care, the high-index states reported higher CHE of 46.1%) than low-index states of 46.7%. In states with a high hospital index, outpatient CHE was 38.4% compared to 37.7% in low-index states, but inpatient CHE is lower in high-index states (44.5%) than in low-index ones (50.6%). Households from states with prior insurance experience show slightly higher outpatient CHE (38.4%) compared to those without (37.6%), whereas for inpatient CHE, households with prior experience report lower CHE (44.5%) compared to those without (51.6%) (Table 3).

Out-of-pocket expenditure (OOPE) for outpatient and inpatient care shows notable differences based on the state health index, district hospital index, and prior insurance experience. For outpatient care, OOPE in states with a high health index is INR 1,730 compared to low-index states of INR 1,736, while for inpatient care, OOPE is substantially higher in high-index states (INR 38,410) than in low-index states (INR 30,680). States with a high hospital index report higher outpatient OOPE (INR 1,771) and inpatient OOPE (INR 35,405) than low-index states with outpatient OOPE of INR 1,640 and inpatient OOPE of INR 32,293. Households with prior health insurance experience more outpatient OOPE (INR 1,771) than those without prior insurance (INR 1,640). Inpatient OOPE is higher for those from states with prior to 2017 health insurance experience (INR 34,823) than for those with no health insurance experience (INR 33,511) (Table 3).

### Regression analysis

**Model 1**, The logistic regression analysis of health insurance coverage, highlights that households living in states with high state health index scores are significantly more likely to have health insurance (AOR 1.069, p < 0.01) (Table 4 showcases results of key variables of the study. For full model see Supplementary Table 2). Similarly, residents in states with a high district hospital index are more likely to be insured (AOR 1.064, p < 0.01). Furthermore, states with health insurance experience before 2018 exhibit a markedly higher likelihood of obtaining coverage for their residents (AOR 6.174, p < 0.01). Additional findings reveal that urban residents are less likely to be insured than rural residents, while larger household sizes are positively associated with insurance coverage. Socio-economic factors, such as higher expenditure deciles and the education level of the household head, particularly at the graduate level, also significantly predict better health insurance coverage (See Supplementary Table 2 for results of full models).

**Model 2**, the logistic regression analysis of outpatient care use, reveals significant associations with the PFHI coverage (AOR 1.291, p < 0.01) (Table 4). The state health index is not significantly associated with outpatient use. Other covariates show that urban residents are less likely to use outpatient services than rural residents, and larger households exhibit higher outpatient care utilisation. Socioeconomic factors, such as higher expenditure deciles, consistently predict greater use of outpatient care. At the same time, the education of the household head shows mixed effects, with primary education increasing utilisation and higher levels of education showing less consistent associations.

**Model 3,** the logistic regression analysis of inpatient care use, indicates that households from states with high district hospital index or having PFHI coverage are not significantly associated. However, the interaction term between PFHI and the district hospital index is significant (1.164, p<0.01). This finding, with the insignificant association of PFHI with inpatient use, indicates that PFHI does better where the district hospital index is higher in delivering higher inpatient utilisation. Prior health insurance experience before 2017 shows a positive association (AOR 1.066, p<0.05) (Table 4). Other covariates reveal that urban residents are less likely to use inpatient care, while larger household sizes are strongly associated with higher utilisation.

**Model 4**, CHE during outpatient care through logistic regression, revealed significant associations with various factors. Publicly funded health insurance (PFHI) was positively associated with increased CHE (AOR 1.097, p<0.01), while other types of insurance showed a negative relationship (AOR 0.920, p<0.1) (Table 5 shows results of key variables. For full model see Supplementary Table 3). state health index was negatively associated weakly (0.958, p<0.1). Urban residents were less likely to experience CHE (AOR 0.910, p<0.01), and larger households had a stronger negative association. Scheduled Tribes and Scheduled Castes demonstrated significantly lower CHE. Higher-income deciles were linked to reduced CHE, with the richest households (10^th^ decile) exhibiting the most substantial negative association (AOR 0.433, p<0.01). Additionally, the education level of the household head had a mitigating effect, as primary education was associated with lower CHE.

**Model 5**, CHE during inpatient care using logistic regression, revealed significant findings related to insurance, health systems, and interaction terms. PFHI was negatively associated with CHE during inpatient care (AOR 0.692 p<0.01), and other forms of insurance also exhibited a negative relationship (AOR 0.631, p<0.01) (Table 5). A high district hospital index or state with health insurance experience before 2017 showed no association with CHE. However, the interaction between PFHI and the district hospital index demonstrated a significant negative association with CHE (AOR 0.883, p<0.01), indicating that PFHI reduces CHE during inpatient care where the district hospital index is higher. The results of the other covariates are almost similar to the findings of Model 4. These findings emphasise the role of PFHI and higher district hospital index in reducing CHE during inpatient care.

**Model 6,** examining log-transformed out-of-pocket expenditure (OOPE) during outpatient care through linear regression, highlighted significant associations with insurance and health system variables. PFHI and other insurance types showed no significant association. A high state health index had a significant negative association with log OOPE (coefficient= −0.100, p<0.01) (Table 5). Urban residents, scheduled tribes and scheduled castes were associated with reduced log-transformed OOPE, while households of larger size, higher expenditure deciles, and heads with graduate education were positively associated. These findings outline key relationships between insurance types, health system indices, and OOPE during outpatient care.

**Model 7**, which examines log-transformed OOPE during inpatient care, finds PFHI had a strong negative association with log-transformed OOPE (coefficient= −0.543, p<0.01). At the same time, other types of insurance showed an even more significant negative coefficient (coefficient= −1.066, p<0.01) (Table 5). A high district hospital index was strongly and negatively associated with OOPE (−0.297, p>0.01). Prior experience with health insurance before 2017 was not associated significantly. The interaction term between PFHI and the district hospital index demonstrated a significant negative association with log-transformed out-of-pocket expenditure (OOPE) (coefficient= −0.252, p<0.01). These results suggest that PFHI and the district hospital index had a substantial mitigating impact on OOPE during inpatient care.

## 4. Discussion

The study findings related to the three questions posed earlier are as follows: better state primary health care is associated with higher PFHI enrollments and a weak association with reduced CHE during outpatient care. In combination with PFHI, better district hospitals are associated with higher use of inpatient care and reduced CHE and OOPE during inpatient care. On their own, a higher district hospital index reduces inpatient OOPE. Prior experience implementing PFHI is associated with higher health insurance coverage and inpatient care use.

### Insurance coverage

In line with a previous study, states with higher state health index were significantly more likely to achieve higher insurance coverage. (25) Similarly, states with high district hospital indexes also showed higher PFHI enrollment. States with a prior history of PFHI implementation exhibited higher PFHI coverage, highlighting the need for long-term commitment and investment in PFHI schemes, which aligns with previous studies on NRHM, where impacts are seen after full implementation. (21)

### Health care use

The district hospital index is positively associated with inpatient care use, which aligns with previous studies. (16,17,27) The significant interaction term between PFHI and the district hospital index further supports these findings, revealing a significant positive effect on inpatient care utilisation. These results suggest that states with robust district hospital systems improve PFHI’s ability to improve inpatient use.

There is a significant positive association between PFHI and outpatient care use, which is surprising as PFHI does not cover outpatient costs. This could be due to increased awareness, improved access to healthcare facilities, and perceived financial protection from implementing the PFHI scheme. This needs further investigation, which is of policy significance, as PFHI is also associated with higher outpatient CHE.

### Financial protection

The study’s most important finding is that the PFHI’s financial protection outcomes are likely to be better for inpatient care in states with better district hospital infrastructure. The interaction term between PFHI and the district hospital index further reduced inpatient CHE (AOR 0.883, p < 0.05), reinforcing the importance of integrating PFHI with robust district hospital systems. The interaction term between PFHI and the district hospital index also reduced inpatient OOPE (coefficient = −0.252, p < 0.01). This underscores the synergistic role of better district hospitals in enhancing PFHI’s effectiveness in reducing OOPE and CHE. State health index was associated with reduced outpatient OOPE (−0.100, p<0.01). The study results show the protective role of PFHI against CHE and OOPE during inpatient care.

Financial protection remains a critical challenge despite PFHI implementation. PFHI was associated with a slight increase in CHE for outpatient care. Other types of insurance and the state health index showed a protective effect, although weakly (p<0.1). Further studies are required to investigate the association of PFHI with higher outpatient use and higher CHE during outpatient care. One hypothesis is that PFHI encourages households to seek care. However, many may be treated as outpatient cases, especially where informal payments are expected for admissions into inpatient care, shifting the financial burden back to households. (28) This could be a more significant problem where households access secondary hospitals directly, skipping primary health care, as often happens in India. (29)

While this study provides important insights, it has some limitations. First, the methodology of relying on the state health index as a proxy for health system strength has been critiqued. (26) The index may not fully capture the quality and accessibility of primary healthcare, potentially influencing the strength of associations observed in the analysis. Second, the cross-sectional nature of the data limits causal inference, making it challenging to determine whether strong health systems directly enhance PFHI outcomes or vice versa. The significant interaction term between PFHI and the district hospital index mitigates this limitation somewhat. Future research should address these gaps.

The findings underscore critical policy implications for advancing UHC in India and other developing countries with similar health systems. Strengthening primary health care and district hospital infrastructure amplifies the impact of PFHI schemes, particularly in reducing inpatient CHE and OOPE. (18,30–32) Primary health care prevents illness and death, leads to a more equitable distribution of health, reduces differences in health across population subgroups, and significantly lowers overall health expenditures. (27,33–35) State governments can take several steps to strengthen health systems: ensure integrated primary health care, allocate adequate financial resources, strengthen health governance, establish meaningful public-private engagement, use digital health tools, deliver innovative services, focus on patients and communities, and adopt new communication technologies. (36,37) Targeted investments in strengthening health systems, especially in low-performing states, can bridge disparities, as evidenced by the significant interaction effects between health systems and PFHI.

The government of India is boosting investments in primary health care. Ayushman Bharat Arogya Mandirs (ABAM) provide free and universal access to services delivered close to communities, including preventive, promotive, curative, and rehabilitative care. Improving the effectiveness of ABAMs has the potential to reduce the financial burden of outpatient care. The study reminds us that more work is required to reduce the financial burden of outpatient care.

Previous studies have highlighted the need to expand coverage, raise awareness about procedures to avail benefits, increase coverage limits, and consider multiple morbidities to reduce inpatient OOPE. (18,30,38) Some have argued for including outpatient care and preventive services in PFHI. (18,30) Expanding PFHI coverage to include outpatient services may be examined to address persistent financial vulnerabilities in outpatient care.

Previous studies have found very high OOPE in private hospitals and an increasing use of private hospitals responsible for CHE. (39,40) Systematic reviews from low- and middle-income countries do not support the claim that the private sector is more efficient, accountable, or medically effective than the public sector; however, the public sector frequently lacks timeliness and hospitality toward patients. (41) Some studies from India have found that private providers, including informal ones, are associated with better completion of checklists and treatment. (21,22) Private speciality hospitals in India are thought to provide good quality care and have unused capacity, in contrast to public hospitals, which are often overcrowded and, as a result, provide poor quality of care.(42) However, due to their high costs, low-income people would not have had access to private hospitals without health insurance. (42)

Studies have supported the strengthening of district hospitals and advocated for building a second district hospital in every district of India. (32) If adequately financed and managed, public hospitals can serve as benchmarks for quality and cost standards across the healthcare system. (43) PFHI can improve the quality of care beneficiaries receive by increasing access to high-quality private hospitals with effective government health insurance programs monitoring the appropriate use, costs, and health outcomes of private hospitals. (42) Strong public and well-regulated private hospitals can lead to higher-quality, cost-effective care.

### Conclusion

This study supports the role of strong health systems in enhancing PFHI schemes’ outcomes. The study results underscore the need for a coordinated approach that aligns PFHI implementation with health system strengthening, ensuring equitable and sustainable healthcare access for all. Moving forward, aligning PFHI design with primary health care and district hospital improvements will be instrumental in addressing India’s healthcare disparities and achieving UHC. A balanced approach— one that strengthens public-sector capacity while integrating private providers under robust governance—appears essential for developing countries to realise the twin goals of UHC and financial protection.

## Data Availability

All data produced are available online at https://microdata.gov.in/NADA43/index.php/catalog/220

## Footnotes

11 The index is calculated by summing standardised scores of the following 12 indicators, equally weighted, for which data at the hospital level is available in the report: hospital beds; doctors, nurses, and paramedical staff availability; availability of support staff, core health services, diagnostic testing services; bed occupancy; C-section rate; surgical productivity; OPD per doctor; and blook bank replacement rate. (2)

2 Also see Supplementary Table 1 for details about all the covariates.

