## Supplemental Tables 1-3 for "Health Systems Disparities and Publicly Funded Health Insurance Outcomes in India: Insights from Comprehensive Annual Modular Survey, 2022-23"

**SUPPLEMENTARY TABLES**

**Supplementary Table 1: Sample distribution and bivariate estimates of outcome by study covariates**

| **Variables** | **Sample distribution** | **Households with any health insurance coverage (%)** | **Households who used outpatient care in the past 30 days (%)** | **Households who used inpatient care for past 365 days (%)** | **Households with Catastrophic health expenditure during outpatient care (%)** | **Households with catastrophic health expenditure during inpatient care (%)** | **Out-of-pocket expenditure among households who sought outpatient care (Indian Rupees)** | **Out-of-pocket expenditure among households who sought inpatient care (Indian Rupees)** |
| --- | --- | --- | --- | --- | --- | --- | --- | --- |
| **State health index** | |  |  |  |  |  |  |  |
| Low | 32.5 | 41.8 | 32.7 | 13.5 | 38.9 | 46.7 | 1,736 | 30,680 |
| High | 67.5 | 68.7 | 32.8 | 12.9 | 37.9 | 46.1 | 1,730 | 38,410 |
| **District hospital index** | |  |  |  |  |  |  |  |
| Low | 30.1 | 36.0 | 32.4 | 12.8 | 37.7 | 50.6 | 1,640 | 32,293 |
| High | 67.5 | 70.2 | 32.4 | 13.2 | 38.4 | 44.5 | 1,771 | 35,405 |
| **Health Insurance experience before 2018** | |  |  |  |  |  |  |  |
| No | 26.0 | 27.3 | 32.8 | 12.8 | 37.6 | 51.6 | 1,660 | 33,511 |
| Yes | 74.0 | 71.4 | 32.3 | 13.2 | 38.4 | 44.5 | 1,757 | 34,823 |
| **Residence** |  |  |  |  |  |  |  |  |
| Rural | 67.1 | 59.3 | 33.2 | 13.5 | 39.5 | 46.8 | 1,623 | 30,688 |
| Urban | 32.9 | 61.3 | 30.7 | 12.3 | 35.5 | 45.2 | 1,972 | 42,970 |
| **Size of the household** | |  |  |  |  |  |  |  |
| 1-3 | 37.3 | 60.6 | 29.0 | 9.1 | 45.9 | 53.1 | 1,573 | 35,875 |
| 4-5 | 42.5 | 61.6 | 32.4 | 13.0 | 35.0 | 46.4 | 1,703 | 34,038 |
| >5 | 20.2 | 55.0 | 38.8 | 20.5 | 33.3 | 40.5 | 2,000 | 33,949 |
| **Religion** |  |  |  |  |  |  |  |  |
| Hindu | 82.9 | 60.7 | 31.3 | 12.7 | 38.4 | 46.8 | 1,734 | 34,962 |
| Islam | 12.0 | 53.9 | 38.1 | 14.7 | 37.0 | 42.6 | 1,579 | 29,767 |
| Christian | 2.6 | 71.9 | 33.4 | 14.6 | 35.9 | 42.7 | 1,872 | 33,374 |
| Others | 2.6 | 50.1 | 39.6 | 15.0 | 40.9 | 51.6 | 2,242 | 44,234 |
| **Social Group** |  |  |  |  |  |  |  |  |
| Others | 25.3 | 61.8 | 34.9 | 13.5 | 40.1 | 48.5 | 2,083 | 43,160 |
| Scheduled tribes | 8.9 | 69.4 | 22.0 | 11.0 | 31.3 | 31.0 | 1,134 | 16,400 |
| Scheduled castes | 20.5 | 59.8 | 33.1 | 13.5 | 38.1 | 44.0 | 1,556 | 27,744 |
| Other backward castes | 45.3 | 57.1 | 32.8 | 13.1 | 38.1 | 48.6 | 1,682 | 35,631 |
| **Expenditure Decile** | |  |  |  |  |  |  |  |
| Poorest Decile | 10.0 | 47.1 | 27.9 | 13.9 | 42.2 | 46.6 | 1,214 | 18,759 |
| 2^nd^ decile | 10.0 | 52.6 | 30.3 | 14.3 | 42.1 | 44.4 | 1,371 | 24,238 |
| 3^rd^ decile | 10.0 | 55.8 | 31.7 | 13.6 | 38.6 | 46.2 | 1,454 | 25,274 |
| 4^th^ decile | 10.0 | 57.5 | 33.0 | 14.3 | 38.8 | 45.7 | 1,546 | 27,180 |
| 5^th^ decile | 10.0 | 58.6 | 33.8 | 13.2 | 37.0 | 47.3 | 1,565 | 30,739 |
| 6^th^ decile | 10.0 | 61.5 | 34.6 | 13.3 | 37.4 | 48.1 | 1,629 | 35,129 |
| 7^th^ decile | 10.0 | 64.4 | 35.4 | 13.4 | 37.9 | 47.2 | 1,728 | 36,213 |
| 8^th^ decile | 10.0 | 65.4 | 34.9 | 12.9 | 38.1 | 47.4 | 1,885 | 44,585 |
| 9^th^ decile | 10.0 | 66.7 | 33.9 | 11.7 | 36.7 | 46.4 | 2,138 | 47,395 |
| Richest Decile | 10.0 | 69.8 | 28.8 | 10.3 | 34.0 | 43.0 | 2,789 | 66,594 |
| **Education of the Head of the Household** | |  |  |  |  |  |  |  |
| Illiterate or < Primary | 24.3 | 56.9 | 32.6 | 13.1 | 39.9 | 45.8 | 1,485 | 27,876 |
| Primary | 16.6 | 62.3 | 34.3 | 13.7 | 36.5 | 43.7 | 1,567 | 31,196 |
| Middle | 15.7 | 58.3 | 32.7 | 13.0 | 36.7 | 44.9 | 1,622 | 30,334 |
| Secondary/ Sn Secondary | 29.3 | 58.2 | 32.7 | 13.6 | 38.7 | 48.3 | 1,871 | 37,966 |
| Graduate or above | 14.2 | 67.6 | 29.0 | 11.4 | 38.1 | 47.7 | 2,245 | 48,918 |

**Supplementary Table 2: Logistic regression of (a) Health insurance coverage (Model 1), (b) Use of outpatient care (Model 2), and (c) Use of inpatient care (Model 3)**

|  | **Model 1**  **Health insurance coverage** | | | **Model 2**  **Use of outpatient care** | | | **Model 3**  **Use of inpatient care** | | |
| --- | --- | --- | --- | --- | --- | --- | --- | --- | --- |
| **Variables** | **AOR** |  | **SE** | **AOR** |  | **SE** | **AOR** |  | **SE** |
| **Insurance (None)** |  |  |  |  |  |  |  |  |  |
| PFHI |  |  |  | 1.291 | *** | 0.017 | 0.995 |  | 0.027 |
| Other insurance |  |  |  | 1.040 |  | 0.027 | 1.093 | *** | 0.031 |
| **Health Systems** |  |  |  |  |  |  |  |  |  |
| State health index (Low) | 1.069 | *** | 0.017 | 1.016 |  | 0.014 |  |  |  |
| district hospital index (Low) | 1.064 | *** | 0.025 |  |  |  | 1.025 |  | 0.033 |
| Before 2018, health insurance experience (No) | 6.174 | *** | 0.187 |  |  |  | 1.066 | ** | 0.030 |
| **Interaction** |  |  |  |  |  |  |  |  |  |
| PFHI*district hospital index |  |  |  |  |  |  | 1.164 | *** | 0.037 |
| **Residence (Rural)** |  |  |  |  |  |  |  |  |  |
| Urban | 0.725 | *** | 0.012 | 0.851 | *** | 0.013 | 0.959 | ** | 0.016 |
| **Household size (1-3)** |  |  |  |  |  |  |  |  |  |
| 4-5 | 1.404 | *** | 0.022 | 1.205 | *** | 0.018 | 1.493 | *** | 0.026 |
| >5 | 1.429 | *** | 0.026 | 1.715 | *** | 0.030 | 2.694 | *** | 0.053 |
| **Religion (Hindu)** |  |  |  |  |  |  |  |  |  |
| Muslim | 0.922 | *** | 0.019 | 1.244 | *** | 0.024 | 1.083 | *** | 0.024 |
| Christian | 1.248 | *** | 0.046 | 1.242 | *** | 0.041 | 1.298 | *** | 0.048 |
| Others | 0.399 | *** | 0.014 | 1.385 | *** | 0.046 | 1.151 | *** | 0.045 |
| **Social Group (Others)** |  |  |  |  |  |  |  |  |  |
| Scheduled Tribes | 1.390 | *** | 0.038 | 0.510 | *** | 0.013 | 0.731 | *** | 0.019 |
| Scheduled Castes | 1.243 | *** | 0.026 | 0.923 | *** | 0.018 | 0.976 |  | 0.021 |
| Other Backward Caste | 1.024 |  | 0.018 | 0.902 | *** | 0.014 | 0.940 | *** | 0.017 |
| **Expenditure Decile (Poorest)** |  |  |  |  |  |  |  |  |  |
| 2^nd^ decile | 1.229 | *** | 0.031 | 1.112 | *** | 0.029 | 1.061 | ** | 0.031 |
| 3^rd^ decile | 1.330 | *** | 0.034 | 1.203 | *** | 0.032 | 1.031 |  | 0.030 |
| 4^th^ decile | 1.314 | *** | 0.033 | 1.291 | *** | 0.034 | 1.108 | *** | 0.032 |
| 5^th^ decile | 1.286 | *** | 0.034 | 1.366 | *** | 0.036 | 1.045 |  | 0.031 |
| 6^th^ decile | 1.406 | *** | 0.038 | 1.444 | *** | 0.038 | 1.086 | *** | 0.033 |
| 7^th^ decile | 1.566 | *** | 0.045 | 1.527 | *** | 0.042 | 1.119 | *** | 0.036 |
| 8^th^ decile | 1.626 | *** | 0.049 | 1.571 | *** | 0.044 | 1.123 | *** | 0.037 |
| 9^th^ decile | 1.745 | *** | 0.059 | 1.611 | *** | 0.050 | 1.069 | * | 0.038 |
| 10^th^ decile (Richest) | 2.199 | *** | 0.083 | 1.454 | *** | 0.050 | 1.054 |  | 0.042 |
| **Head of household education (Illiterate)** |  |  |  |  |  |  |  |  |  |
| Primary | 1.045 | ** | 0.020 | 1.043 | ** | 0.020 | 1.057 | *** | 0.022 |
| Middle | 0.932 | *** | 0.019 | 0.972 |  | 0.019 | 1.009 |  | 0.022 |
| Secondary/ Sn. Secondary | 0.922 | *** | 0.016 | 0.971 | * | 0.017 | 1.098 | *** | 0.021 |
| Graduate | 1.454 | *** | 0.038 | 0.867 | *** | 0.021 | 1.000 |  | 0.027 |
| **Constant** | 0.290 | *** | 0.010 | 0.348 | *** | 0.012 | 0.086 | *** | 0.003 |
| **Observations** | **3,02,086** | | | **3,02,086** | | | **3,02,086** | | |

*** p<0.01, ** p<0.05, * p<0.1

AOR= adjusted odds ratio, CHE= catastrophic health expenditure, PFHI = publicly funded health insurance (AB-PMJAY or State Health Insurance), OOPE = out-of-pocket expenditure

**Supplementary Table 3: Logistic regression for (a) CHE during outpatient care (Model 4), (b) CHE during inpatient care (Model 5), and linear regression for (c) log OOPE during outpatient care (Model 6), and (d) Log OOPE during inpatient care**

|  | **Model 4**  **CHE during outpatient care (log regression)** | | | **Model 5**  **CHE during inpatient care (log regression)** | | | **Model 6**  **Log OOPE during outpatient care (linear regression)** | | | **Model 7**  **Log OOPE during inpatient care (linear regression** | | |
| --- | --- | --- | --- | --- | --- | --- | --- | --- | --- | --- | --- | --- |
| **Variables** | **AOR** |  | **SE** | **AOR** |  | **SE** | **coef.** |  | **SE** | **coef.** |  | **SE** |
| **Insurance (None)** |  |  |  |  |  |  |  |  |  |  |  |  |
| PFHI | 1.097 | *** | 0.024 | 0.692 | *** | 0.034 | 0.018 |  | 0.018 | -0.543 | *** | 0.059 |
| Other insurance | 0.920 | * | 0.041 | 0.631 | *** | 0.033 | -0.048 |  | 0.033 | -1.066 | *** | 0.075 |
| **Health Systems** |  |  |  |  |  |  |  |  |  |  |  |  |
| State health index (Low) | 0.958 | * | 0.022 |  |  |  | -0.100 | *** | 0.019 |  |  |  |
| district hospital index (Low) |  |  |  | 0.962 |  | 0.058 |  |  |  | -0.297 | *** | 0.074 |
| Before 2018, health insurance experience (No) |  |  |  | 0.982 |  | 0.053 |  |  |  | -0.089 |  | 0.068 |
| **Interaction** |  |  |  |  |  |  |  |  |  |  |  |  |
| PFHI*district hospital index |  |  |  | 0.883 | ** | 0.051 |  |  |  | -0.252 | *** | 0.073 |
| **Residence (Rural)** |  |  |  |  |  |  |  |  |  |  |  |  |
| Urban | 0.910 | *** | 0.023 | 0.924 | *** | 0.027 | -0.088 | *** | 0.021 | -0.269 | *** | 0.042 |
| **Household size (1-3)** |  |  |  |  |  |  |  |  |  |  |  |  |
| 4-5 | 0.569 | *** | 0.014 | 0.716 | *** | 0.022 | 0.335 | *** | 0.021 | 0.258 | *** | 0.043 |
| >5 | 0.486 | *** | 0.014 | 0.537 | *** | 0.018 | 0.666 | *** | 0.023 | 0.412 | *** | 0.048 |
| **Religion (Hindu)** |  |  |  |  |  |  |  |  |  |  |  |  |
| Muslim | 0.926 | ** | 0.029 | 0.773 | *** | 0.030 | 0.014 |  | 0.023 | -0.084 |  | 0.052 |
| Christian | 0.963 |  | 0.054 | 0.985 |  | 0.066 | -0.181 | *** | 0.057 | 0.091 |  | 0.101 |
| Others | 1.297 | *** | 0.069 | 1.363 | *** | 0.093 | 0.354 | *** | 0.038 | 0.364 | *** | 0.095 |
| **Social Group (Others)** |  |  |  |  |  |  |  |  |  |  |  |  |
| Scheduled Tribes | 0.571 | *** | 0.027 | 0.444 | *** | 0.022 | -0.611 | *** | 0.038 | -1.340 | *** | 0.072 |
| Scheduled Castes | 0.807 | *** | 0.026 | 0.749 | *** | 0.029 | -0.280 | *** | 0.026 | -0.484 | *** | 0.052 |
| Other Backward Caste | 0.875 | *** | 0.022 | 0.962 |  | 0.030 | -0.166 | *** | 0.020 | -0.160 | *** | 0.042 |
| **Expenditure Decile (Poorest)** |  |  |  |  |  |  |  |  |  |  |  |  |
| 2nd decile | 0.940 |  | 0.042 | 0.896 | ** | 0.047 | 0.243 | *** | 0.035 | 0.190 | *** | 0.067 |
| 3rd decile | 0.774 | *** | 0.034 | 0.932 |  | 0.049 | 0.270 | *** | 0.035 | 0.387 | *** | 0.067 |
| 4th decile | 0.764 | *** | 0.034 | 0.915 | * | 0.047 | 0.384 | *** | 0.035 | 0.616 | *** | 0.065 |
| 5th decile | 0.675 | *** | 0.030 | 0.939 |  | 0.050 | 0.433 | *** | 0.035 | 0.749 | *** | 0.066 |
| 6th decile | 0.670 | *** | 0.030 | 0.962 |  | 0.053 | 0.499 | *** | 0.036 | 0.991 | *** | 0.068 |
| 7th decile | 0.657 | *** | 0.031 | 0.896 | * | 0.051 | 0.577 | *** | 0.039 | 1.014 | *** | 0.073 |
| 8th decile | 0.633 | *** | 0.030 | 0.874 | ** | 0.051 | 0.696 | *** | 0.039 | 1.233 | *** | 0.072 |
| 9th decile | 0.554 | *** | 0.029 | 0.805 | *** | 0.052 | 0.855 | *** | 0.042 | 1.312 | *** | 0.080 |
| 10th decile (Richest) | 0.433 | *** | 0.025 | 0.627 | *** | 0.045 | 0.925 | *** | 0.050 | 1.526 | *** | 0.094 |
| **Head of household education (Illiterate)** |  |  |  |  |  |  |  |  |  |  |  |  |
| Primary | 0.895 | *** | 0.028 | 0.919 | ** | 0.034 | -0.073 | *** | 0.026 | 0.026 |  | 0.053 |
| Middle | 0.934 | ** | 0.031 | 0.933 | * | 0.036 | -0.038 |  | 0.027 | 0.081 |  | 0.052 |
| Secondary/ Sn. Secondary | 1.059 | ** | 0.030 | 1.025 |  | 0.035 | 0.102 | *** | 0.023 | 0.141 | *** | 0.046 |
| Graduate | 1.122 | *** | 0.045 | 1.052 |  | 0.050 | 0.223 | *** | 0.031 | 0.101 |  | 0.069 |
| Constant | 1.728 | *** | 0.095 | 2.499 | *** | 0.173 | 5.985 | *** | 0.044 | 9.032 | *** | 0.088 |
| **Observations** | **90,194** | | | **74,256** | | | **90,194** | | | **74,256** | | |

*** p<0.01, ** p<0.05, * p<0.1

AOR= adjusted odds ratio, CHE= catastrophic health expenditure, PFHI = publicly funded health insurance (AB-PMJAY or State Health Insurance), OOPE = out-of-pocket expenditure.
